# The efficacy and safety of tyrosine kinase inhibitors in advanced and metastatic thyroid cancer: Study Protocol for a systematic review and meta-analysis of Phase III and IV randomised controlled trials

**DOI:** 10.1101/2023.05.07.23289595

**Authors:** Jessica Bindra, Ruan Vlok, Matti Gild

## Abstract

**Background and Aims:** Tyrosine kinase inhibitors (TKIs) are now readily considered in the management of advanced and metastatic thyroid cancer. However, a key limiting factor are their adverse events (AEs), which can impair quality of life and influence patient acceptance of treatment. Prior systematic reviews have assessed the efficacy and safety of TKIs by reviewing both retrospective and prospective studies and further Phase III randomised controlled trials (RCTs) have since been published. We therefore plan to conduct a systematic review and meta-analysis of only prospective phase III and IV RCTs including the most recently published trials to investigate the efficacy and AEs of TKIs in patients with advanced and metastatic thyroid cancer compared to placebo.

**Review methods:** The Cochrane Central Register of Controlled Trials (CENTRAL) in the Cochrane Library, OVID Medline, Embase and PubMed databases will be electronically searched. Grey literature will be searched through Clinicaltrials.gov and www.ISRCTN.com. Expert informants will be contacted. We will include Phase III and IV randomised controlled trials that investigate the efficacy and AEs of TKIs in adults with advanced or metastatic thyroid cancer compared to placebo. Two authors will extract and assess for risk of bias using the Cochrane Risk of Bias Tool. Data extraction will include outcomes for objective response rate (ORR), progression free survival (PFS) and AEs, including diarrhoea, nausea, hypertension, proteinuria, palmar-plantar erythrodysaesthesia (PPE) and grade 3+ AEs, with a subgroup analysis on medullary vs differentiated thyroid cancer, TKI type and history of prior TKI or other systemic therapy. Pooled effect sizes will be meta-analysed using Review Manager 5 software through a random effects model. The study will be reported in line with the PRISMA guidelines.

**Discussion:** This will be the first systematic review to be conducted on the safety and efficacy of TKIs in advanced or metastatic thyroid cancer to meta-analyse phase III and IV RCT level data and include the most recently published trials. This review aims to highlight the balance between the therapeutic effect and toxicity of TKIs, and guide future studies that may be required in exploring different dosing and timing of TKI therapy.

## BACKGROUND

Thyroid cancer can be categorised into medullary thyroid cancer (MTC), anaplastic thyroid cancer (ATC) and differentiated thyroid cancer (DTC), which includes papillary thyroid cancer (PTC) and follicular thyroid cancer (FTC). Radioactive iodine (RAI) is a well-established treatment for metastatic DTC; however, some tumours demonstrate resistance to RAI therapy and carry a poorer prognosis.^1^ Furthermore, RAI cannot be applied to MTC as the cells do not uptake iodine, thereby limiting therapeutic options for metastatic MTC. Tyrosine kinase inhibitors (TKIs) have emerged as a promising option in RAI-refractory DTC and metastatic MTC over the past 10 years.^2^

Many randomised controlled trials (RCTs) have investigated the efficacy and safety of TKIs in patients with advanced or metastatic thyroid cancer, including lenvatinib,^3^ sorafenib,^4^ cabozantinib^5^ and vandetanib,^6^ with demonstration of improvements in overall survival, progression-free survival (PFS) and objective response rate (ORR). TKIs are now readily considered in the management of advanced and metastatic thyroid cancer. However, a key limiting factor are their adverse events (AEs), which can impair quality of life and influence patient acceptance of treatment. Common adverse events include diarrhoea, nausea, hypertension, proteinuria and palmar-plantar erythrodysaesthesia (PPE), which may necessitate dose reduction or cessation. Further research is required into the effects of TKI therapy on patients, as this will guide future studies to investigate optimal dosing regimens and timing of therapy to achieve improved patient outcomes (both clinical and self-reported).

Prior systematic reviews have assessed the efficacy and safety of TKIs by reviewing both retrospective and prospective studies including non-controlled and Phase I/II trials,^2,7-9^ and further Phase III and IV RCTs have since been published. We therefore plan to conduct systematic review and meta-analysis of only prospective phase III and IV RCTs including the most recently published trials to investigate the efficacy and AEs of TKIs in patients with advanced and metastatic thyroid cancer compared to placebo.

## OBJECTIVES

We will perform a systematic review and meta-analysis describing the safety and efficacy of TKIs in radioiodine-refractory (RAI-R) advanced and metastatic thyroid cancer. We will report according to the PRISMA guidelines^10^ and will publish the protocol on a preprint server.^11^ Our primary objective is to describe the efficacy of TKIs, measured by ORR and PFS, and incidence of the most common AEs encountered by patients on TKIs in the published literature. Due to the expected heterogeneity, we aim to assess the association between thyroid cancer type (PTC/FTC, MTC, ATC), TKI type, and the presence of AEs.

## METHODS

### 1. Criteria for considering studies for this review

#### 1A. Types of studies and participants

We will include Phase III and IV randomised controlled trials that investigate the efficacy and AEs of TKIs in adults with advanced or metastatic thyroid cancer compared to placebo. Studies will be required to report the inclusion and exclusion criteria for their patients, TKI treatment and dosing, thyroid cancer stage, definition of prognostic outcomes and AEs. Outcome measures will include ORR, PFS and AEs. Patients must be over 16 years of age. Studies must be published in full text. Studies will be excluded if they are retrospective, single-arm trials, cohort studies, cross-sectional studies and duplicate studies.

#### 1B. Types of interventions

We will include studies that assess any TKI type at any dose for any duration. All thyroid cancer types will be included. In order to be included, a study must report the following:

1. The TKI used;
2. The TKI dose administered; and
3. At least 1 included outcome of interest (ORR, PFS and/or AEs).

### 2. Data collection and analysis

#### 2A. Selection of studies

Trial selection will be conducted independently and in duplicate by two review authors. JB and an additional author will independently screen all titles and abstracts of each reference identified by our search and will then independently assess the full text of any potentially relevant studies for eligibility or exclusion. The reviewers will not be blinded to journal or author names. We will use Covidence software to collate search results, remove duplicates and record screening decisions and exclusions at each stage.^12^ This will be facilitated by a standardised digital screening, inclusion and exclusion tools, in line with methods outlined in the *Cochrane Handbook for Systematic Reviews of Interventions*.^13^ We will resolve any disagreements by discussion and consensus prior to proceeding at each stage. We will record the process in sufficient detail to produce a PRISMA flow diagram.^10^

#### 2B. Data extraction and management

Through Covidence, we will use a standardised digital data-extraction sheet, in line with requirements outlined in the Cochrane Handbook for Systematic Reviews of Interventions Version 6.3.^13^ Two review authors (JB and an additional author) will independently extract information regarding trial design, data regarding the TKI therapy, study and population characteristics and outcomes of interest (reported below in Section 3) and information relevant to Risk of Bias grading. Study characteristics will include design, randomisation method, TKI regimen (dose, duration, and length of follow-up) and reason for withdrawals. Population characteristics will include age, sex, type of thyroid cancer, and previous treatment, including prior TKI therapy. Quality and certainty of evidence will be assessed using the Grades of Recommendation, Assessment, Development and Evaluation (GRADE) approach.^13-15^ Where required, we (JB and an additional author) will contact individual trial authors or organisations to obtain missing data or clarification regarding unclear data. We will resolve any disagreements by discussion and consensus.

#### 2C. Assessment of risk of bias in included studies

Two review authors (JB and an additional author) will independently assess methodological quality using Cochrane’s Risk of Bias 2 (ROB 2) tool.^13^ Domains assessed will include randomisation, allocation concealment, blinding of participants, personnel and outcome assessment, incomplete outcome data, selective reporting and whether an intention-to-treat analysis was performed. We will present a ‘Risk of Bias’ summary, as well as ‘Risk of Bias’ judgements for individual studies in the ‘Characteristics of included studies’ tables.

### 3. Important outcomes

The primary outcome will be PFS, the commonest AEs encountered by patients on TKI therapy (predicted to include nausea, diarrhoea, fatigue, PPE, and hypertension) and the presence of grade 3 or higher adverse events, which is considered to potentially impair daily functioning and affect continuation of treatment. Secondary outcomes will include ORR, driven by complete response (CR) and partial response (PR).

### 4. Search methods for identification of studies

#### 4A. Electronic searches

We will electronically search the Cochrane Central Register of Controlled Trials (CENTRAL) in the Cochrane Library, OVID Medline, Embase and PubMed databases using key words, synonyms and subject headings that relate to thyroid cancer and TKIs. Grey literature will be searched through Clinicaltrials.gov and www.ISRCTN.com. A comprehensive search will be performed on references of retrieved articles, systematic reviews and by asking expert informants. Where required, study authors will be contacted to clarify details and for additional information. No restrictions will be applied to language or publication date.

### 5. Data synthesis

#### 5A. Statistical methods and assessment of heterogeneity

For PFS, results will be presented as a summary hazard ratio (HR) with a 95% confidence interval (CI). For dichotomous data, results will be presented as a summary risk ratio (RR) with a 95% CI. An RR summary estimate has been chosen as the rate of ORR and AEs are expected to be greater than 10% in each trial, based upon clinical experience. Response rates will be evaluated according to the Response to the Evaluation Criteria in Solid Tumours (RECIST) criteria.^16^

We plan to perform analyses using STATA (MP version 16.1, College Station, TX, USA) and the RevMan 5 software.^17^ If feasible with the number of studies identified, we plan to perform a network analysis to compare the different TKI types compared to placebo with regards to the outcomes of interest. Clinical heterogeneity will be addressed by using a random effects model (inverse variance method). Heterogeneity will be assessed amongst trials by using the I^2^ statistic and Chi^2^ test. Strong heterogeneity is defined as an I^2^ of over 50%, and P ≤ 0.10 with the Chi^2^ test. Where more than ten studies are identified, a funnel plot will be interrogated to assess for publication bias. The study will be reported in line with the PRISMA guidelines.

#### 5B. Subgroup analysis

Where sufficient data is available, subgroup analysis will be performed for each outcome based upon:

1. Type of thyroid cancer (DTC, MTC, ATC)
2. Type of TKI
3. Dose of TKI
4. History of prior systemic therapy (particularly TKI therapy)

#### 5C. Sensitivity analysis

A sensitivity analysis will be conducted where possible to test the robustness and reliability of results, particularly considering patients who have previously received prior TKI or other systemic therapy.

#### 5D. Dealing with missing data

When data is found to be missing, trial authors will be contacted to clarify reasons, and include these reasons in our discussion. Analysis will be performed by intention-to-treat principle where possible.

## DISCUSSION

To our knowledge, this will be the first systematic review on the safety and efficacy of TKIs in advanced or metastatic thyroid cancer to meta-analyse phase III and IV RCT level data and include the most recent RCTs published. We aim to highlight the need to balance the therapeutic effect and toxicity of TKIs. We aim to guide future studies to explore the safety and efficacy of starting TKIs at different doses and different timings, depending on the AE findings in this review.

### Intended search strategy in Ovid format

#1 exp thyroid cancer/

#2 exp thyroid tumor/

#3 exp thyroid medullary carcinoma/

#4 exp thyroid follicular carcinoma/

#5 exp thyroid papillary carcinoma/

#6 exp anaplastic thyroid carcinoma/

#7 exp differentiated thyroid cancer/

#8 exp thyroid gland/

#9 exp malignant neoplasm/

#10 (#8 and #9)

#11 (#1 or #2 or #3 or #4 or #5 or #6 or #7 or #10)

#12 exp protein tyrosine kinase inhibitor/

#13 exp lenvatinib/

#14 exp sorafenib/

#15 exp vandetanib/

#16 exp cabozantinib/

#17 exp motesanib/

#18 exp rivoceranib/

#19 exp axitinib/

#20 exp sunitinib/

#21 exp selumetinib/

#22 exp pazopanib/

#23 (#12 or #13 or #14 or #15 or #16 or #17 or #18 or #19 or #20 or #21 or #22)

#24 exp randomized controlled trial/

#25 exp controlled clinical trial/

#26 random*.ab.

#27 placebo.ab.

#28 drug therapy.fs.

#29 trial.ab.

#30 groups.ab.

#31 (#24 or #25 or #26 or #27 or #28 or #29 or #30)

#32 exp animals/ not humans.sh.

#33 (#31 NOT #32)

#34 (#11 AND #23 AND #33)

## Data Availability

All data produced in the present work will be contained in the manuscript.

